# Traumatic Brain Injury among football players in 2017 Series A Brazilian Football Championship – The gap between concussion protocols and the current medical practices

**DOI:** 10.1101/2020.05.14.20101212

**Authors:** Cármine Porcelli Salvarani, Lucas Ribeiro de Medeiros, Leonardo Henrique Micheletti Sotocorno, Vinícius Simon Tomazini, Diego Ciotta de Castro, Eduardo Almeida Dias, Paulo Sérgio Teixeira da Costa, Bruno Bueno Pimenta, Diego Almeida de Oliveira, Eduardo Vinícius Colman da Silva, Fernando Henrique Sapatero

**Author notes:** Corresponding Author; E mail.

## Abstract

**Background:** Sideline assessment of football players with a potential concussion is a challenging concern. Video analysis is an important tool to recognize traumatic brain injury (TBI), including sports-related concussions, among football players.

**Aim:** To report the characteristics of TBI among football players in 2017 Brazilian Series A Football Championship and to discuss the evaluation protocols of football players with concussion.

**Methods:** This is an observational study concerning video analysis of all matches on 2017 Series A Brazilian Football Championship. The videos were first analyzed by a team of 10 trained medical students. All suspected TBIs were reviewed by the research’s coordinator. Concussion diagnosis was defined by one of the following events: lying motionless, loss of responsiveness, impact seizure, disorientation and motor incoordination. The length of sideline medical assessment was systematically recorded.

**Results:** In a whole of 380 matches, it was verified 374 TBIs. The most common etiology was “head-to-head” impact. Twelve players were replaced after TBI. Thirteen players (3,5% of TBIs) had concussion, but only four of them were replaced. The overall mean time for medical assessment was 1’35” (range: 48” to 7’40”). Considering the twelve players who had a concussion and the thirteen players who were replaced, the mean time for medical evaluation was 3’19” and 3’26”, respectively.

**Conclusion:** This study confirms that there is a gap between formal protocols for concussion assessment on sideline in Brazilian Elite Football and current medical practices. It also discusses changes on football rules of player replacement to an adequate sideline medical evaluation.

## INTRODUCTION

Sport-related concussion (SRC) is currently a worldwide critical public health concern. Concussion is a subset of mild Traumatic Brain Injury (TBI) and it is defined as traumatically induced transient disturbance of brain function that involves a complex pathophysiological process. Based on acute injury characteristics, concussion is placed at the less severe end of the brain injury spectrum.[1]

In the world of sports, the highest incidence of SRC occurs in American football, hockey, rugby, football and basketball. In Football, head injury can be a result of contact of the head with another head (or other body parts), ground, goal post, other unknown objects or even an unexpected ball hit. By the global popularity of football, the overall contribution to total SRC is likely to significantly over other sports.[2]

According to recent publications, the neurological understanding of SRC consequences and its effect in athletics life, especially regarding football, is still not properly done. As a result, development of effective tools for proper management of these injuries has been reported by current research. The sideline assessment of SRC in football is challenging due to the variability of presentations, the specific environment, the reliance on athlete-reported symptoms and the varying specificity and sensitivity values of sideline assessment tools. The correct assessment of suspected concussion immediately after injury is an important practice for early diagnosis, appropriate management of athletes with TBI and even for players’ safe return to the match.[3,4]

There are recommended protocols provided by the consecutives Third, Fourth and Fifth International Conferences on Concussion in Sports when an athlete shows any sign of concussion.[5–7] The lasts Sport Concussion Assessment Tools (SCAT) developed by Consensus Statement on Concussion in Sports were adopted by Fédération Internationale de Football Association (FIFA).[8,9] However, previous reports using video analysis of 2014 Men’s FIFA world cup and 2016 Men’s Union of European Football Association (UEFA) found that a potential concussion event was not correctly evaluated, showing a lack of congruence between these protocols and practices on field.[10,11]

This study is the first specific report with video analysis of TBI among football players in Brazilian Series A Football Championship, regarding the complete epidemiology, in order to verify the characteristics of TBI in Brazilian Elite football and to discuss about the medical protocols for concussion in football matches.

## METHOD

This is an observational study concerning video analysis of all 380 matches (38 rounds, 10 matches each round) of 2017 Series A Brazilian Football Championship, based on recent publications related to SRC with video analysis.[10–14] The videos with suspected TBI were first analyzed at the same time the matches took place by a team of 10 trained medical students of State University of Maringá (PR/Brazil), under the coordination of a Neurosurgery professor. After first analysis, all suspected TBIs were reviewed by the research coordinator.

### 1. Guideline and coding of events

A guideline was built to record data about TBI characteristics. In order to fulfill the guideline, TBI was considered as any event in which one or more football players had a head impact injury (head x head, head x ball or head x elbow for instances). At many times, Brazilian football players claimed a TBI, but were, in fact, pretending to gain advantages in the match. Besides the videos, the television narrations were taken to add some information about TBI. It was considered that a “head-to-head” TBI as a double TBI, that means, two TBIs at the same moment. All videos with suspected TBI were inserted in an application smartphone platform (WhatsApp^R^) and shared among all student’s team and the research coordinator.

For concussion diagnosis, it was defined one of the following events: lying motionless, loss of responsiveness, impact seizure, disorientation and motor incoordination. When a football player was replaced due to TBI, the after match medical information was taken by media information and/or official football team medical statement.

### 2. Medical Assessment

The length of sideline medical assessment was systematically recorded in all cases of TBI that required a medical attendance.

### 3. Patient and public involvement

Patients and/or public were not involved in this research.

## RESULTS

In a whole of 380 matches, it was verified 374 TBIs, almost 1 TBI by match (0,98). There were 30 cut blunt injuries (CBI) among TBIs and 3 players were replaced to have wounds sutured due to the extension of CBI. The other 27 players were treated by compressive dressing with swimming caps and were not replaced. The overall findings, with etiology, CBI, concussion and players’ replacement are shown on Table 1.

**Table 1.** Characteristics of Traumatic Brain Injury concerning overall events, relation with matches, etiology, cut blunt injury, concussion and players replacement.

|  |  |  |  |
| --- | --- | --- | --- |
| **Total of Traumatic Brain Injuries (TBI)** | **374** | **Total of matches** | **380** |
| **TBI by match** | **0,98** | **Matches with TBI** | **195** |
| **TBI by round** | **9,98** | **Matches without TBI** | **185** |
| **Etiology** |  | **Cut blunt wound** | **30** |
| **head x head (double TBI)** | 91 (182) | head x head | 19 |
| head x elbow | 86 | Head x elbow | 6 |
| head x foot | 29 | head x foot | 4 |
| head x unexpected ball hit | 17 | head x knee | 1 |
| head x knee | 12 |  |  |
| head x leg | 11 | **Player replacement after TBI** | 12 |
| head x shoulder | 10 |  |  |
| head x hip | 9 | **Concussion (3,5% of all TBI)** | 13 |
| head x hand | 7 |  |  |
| head x floor | 4 | **Player replacement after concussion** | 4 |
| head x others | 7 |  |  |

Thirteen players had concussion (3,5% of all TBIs) and only four of them were replaced. Among these players who had concussion, there were 2 players who were assisted by Intensive Care Unit in the field. These 2 players had impact seizure and, in both situations, were removed to a hospital (Figure 1). The post-match Computed Tomography and MRI of both players were normal.

**Figure 1.**
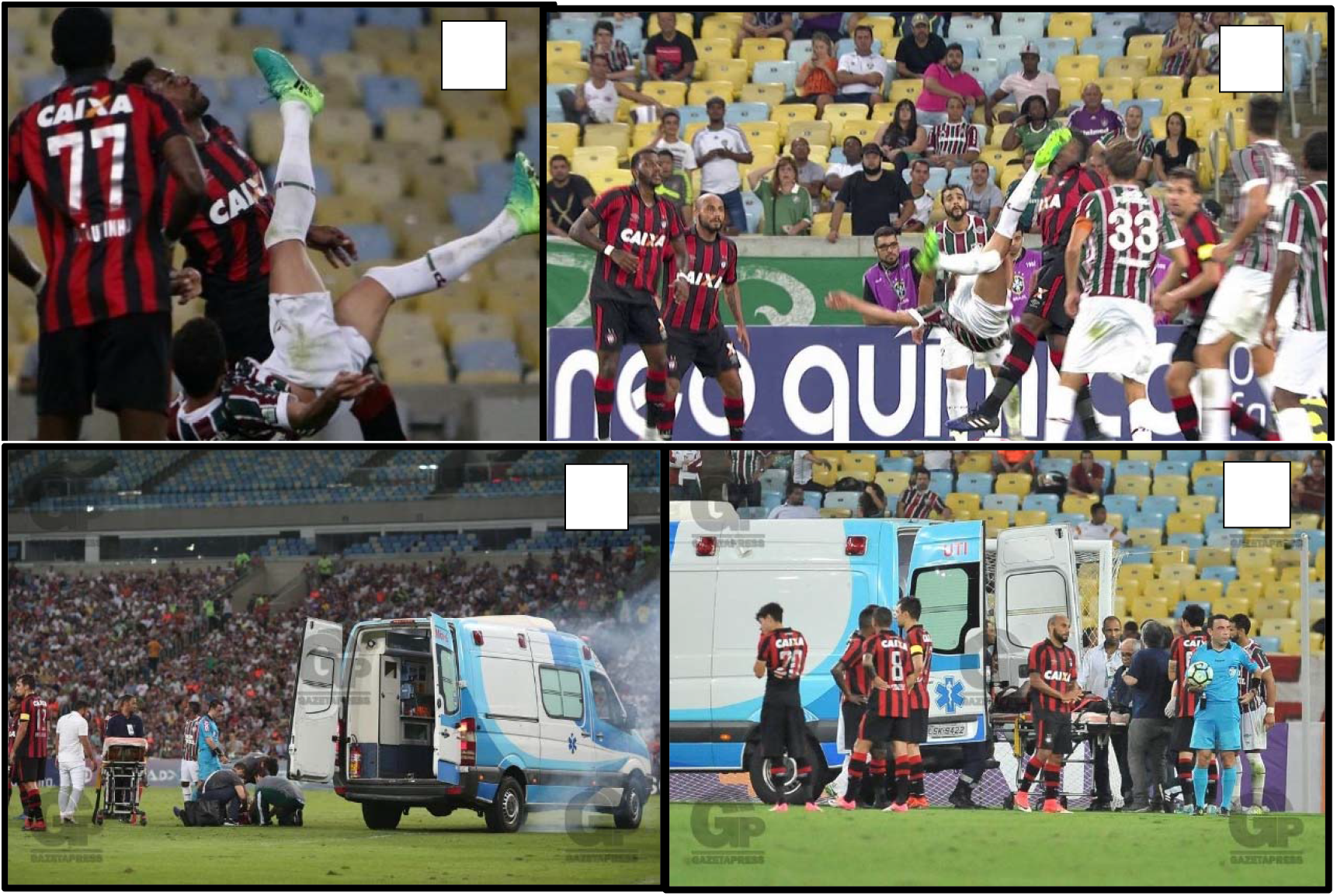
A, B. Player’s photo showing TBI with later impact seizure, C, D. Intensive Care Unit on the field during medical assessment to remove the player to a hospital

The maximum number of TBIs in one match was 7, but no players were replaced. Other particular fact was that 3 TBIs occurred in just one play involving 3 players (triple head TBI).

There were 82 events of TBI without medical assessment required. In general, the mean time for medical assessment among remaining 292 TBIs was 1’35” (from 48” to 7’40”). Considering the 13 players who had concussion and the 12 players who were replaced, the mean time for medical evaluation was 3’26” and 3’19”, respectively (see Table 3 and 4). If the two reported severe TBIs, which an intensive care unit ambulance was in field, were excluded from the average, the mean time for medical evaluation goes down to 2’43” for players with concussion and 2’54” for replaced players after TBI. In both situations, length of medical evaluation was very similar despite the occurrence of concussion. Comparing between the overall mean time for medical assessment and player with concussion, there was an increase of 1’51”.

The main characteristics of players with concussion and replaced players after TBI are described on Table 2 and 3.

**Table 2.** Relationship between etiology of concussion, player replacements and length of medical assessment after TBI.

| **Etiology** | **Number of concussion** | **Player replacement after concussion** | **Time for medical evaluation (mean time: 3'26"/2'43" \*)** | **Observations** |
| --- | --- | --- | --- | --- |
| <u>Unexpected ball hit x head</u> | 4 | 0 | Case 1: 2'40"<br>Case 2: 2'30"<br>Case 3: 2'50"<br>Case 4: 2'42" |  |
| <u>Elbow x head</u> | 3 | 1 | Case 5: 2'30"<br>Case 6: 2'45"<br>Case 7: 2'50" | Maintained disorientation |
| <u>Foot x head</u> | 2 | 2 | Case 8: 2'50"<br>Case 9: 7'40" ** | Blunt cut injury associated<br>Player removed to hospital |
| <u>Knee x head</u> | 2 | 1 | Case 10: 7'10" ** | Player removed to hospital |
|  |  |  | Case 11: 2'45" |  |
| <u>Head x head</u> | 1 | 0 | Case 12: 2'50" |  |
| <u>Hip x head</u> | 1 | 0 | Case 13: 2'40" |  |
| **Total** | **13** | **4** |  |  |

\* Mean time for medical evaluation without cases 9 and 10
\*\* In both situations, players were removed from field by intensive care unit ambulance to hospital

**Table 3.** Relationship between etiology, causes of player replacements (concussion and non-concussion events) and length of medical assessment after TBI.

| **Etiology** | **N** | **Cause/Observation** | **Time for medical evaluation (mean time: 3'19"/2'54" \*)** |
| --- | --- | --- | --- |
| Knee x head | 1 | Case 1: impact seizure/player removed to hospital | 7'10" ** |
| Foot x head | 2 | Case 2: impact seizure/player remove to hospital | 7'40" ** |
|  |  | Case 3: concussion/blunt cut injury associated | 2'50" |
| Head x head | 4 | Case 4: sub galeal hematoma and dizziness | 2'00" |
|  |  | Case 5: blunt cut injury | 5'30" |
|  |  | Case 6: dizziness | 5'00" |
|  |  | Case 7: blunt cut injury | 2'50" |
| Elbow x head | 4 | Case 8: concussion/maintained disorientation | 2'30" |
|  |  | Case 9: blunt cut injury | 1'45" |
|  |  | Case 10: blunt cut injury | 1'40" |
|  |  | Case 11: headache | 2'20" |
| Shoulder x head | 1 | Case 12: headache | 2'40" |
| Total of replacements | 12 |  |  |

\* Mean time for medical evaluation without cases 9 and 10
\*\* In both situations, players were removed from field by intensive care unit ambulance to hospital

## DISCUSSION

There are unforgettable TBIs reports along the history of football: Leonardo and Tab Ramos, in a match between Brazil and United States of America at the 1994 FIFA World Cup; Petr Cech, goal keeper of Chelsea, during a match against Reading in 2006 Premier League Championship, Fernando Torres in 2017 at La Liga Championship. In Brazil, a football player called “Vagner Bacharel” died 6 days after a TBI during a match on 1990. The misdiagnosed acute epidural hematoma was the etiology of his death. Year after year, TBI among football players has been growing and the concern with their assessment has been reported.[15–18]

Despite the limitations of video analysis that doesn’t provide full information of TBI and medical assessment, it may be a useful adjunct to the sideline assessment of a possible concussion.[16] This tool was used in recent studies on 2014 Men’s FIFA World Cup and 2016 Union of European Football Association (UEFA) European Championship.^10 11^ According to these recent publications, the results of this first study among soccer players in Brazil, supported by video analysis, showed how the concussion was evaluated on sideline. Besides, it was studied the overall epidemiologic data of TBI, the length of medical assessment of players who had concussion and/or were replacement after TBI.

In terms of football biomechanics, head-to-head impacts and unexpected ball hits with great speed result in the highest head acceleration and, consequently, higher risks of concussion.[2,19] This is important because, in our findings, the most frequent TBI event was head-to-head (91 events resulting 182 TBIs), however there were 4 concussions after an unexpected ball hit (in a total of 17 events) and only one case of concussion after head-to-head impact.

Updated protocols for sideline assessment of an athlete with a concussion were published following the International Conferences on Concussion in Sport on 2008, 2012 and 2016.[5–7] The Concussion in Sports Group recommendations assert that when an athlete shows any features of a concussion, the athlete should be evaluated by a physician or another licensed healthcare provider onsite, assessed using SCAT or other sideline concussion assessment tools and prevented from Return to Play in the event of a positive diagnosis. The SCAT 5 is the actual assessment tools supported and endorsed by FIFA. Furthermore, according SCAT5 medical evaluation cannot be performed correctly in less than 10 minutes.[9]

On Men’s 2014 FIFA World Cup and on 2016 UEFA European Championship, 63% and 72,5% of athletes with potential concussion event (PCE) were not medically assessed, respectively. All players with PCE that were medically assessed resulted in no replacement after TBI.[10,11]

In this study, only 4 players out of 13 with a concussion during the Championship were replaced. Despite the 2 players who were assisted by an intensive care unit inside the field and removed directed to hospital due to an impact seizure and the 1 player who had an extensive BCI associated with the concussion, just 1 player was replaced after the concussion without any associated injuries. Besides, in two different moments, video images showed that the decision to return to the match was motivated and pressured more by the athlete’s desire than the medical staff itself.

Sport’s physicians should be specifically trained to provide care along the continuum of the concussion, from the acute injury to return-to-play decisions.[2] According to SCAT5 adopted by FIFA,[9] the protocols cannot be performed in less than 10 minutes. During the 2014 FIFA World Cup Championship, the mean time for player assessment after suspected concussion was 1′47” (range: 1′04”-3′).[11] In our findings, in a whole of 374 TBIs, 82 players didn’t receive medical attendance. The mean time for 292 players with TBIs which received assessment was 1′35” (range: 48”-7′40”). Considering only the 13 players who had a concussion and the 12 players who were replaced, the mean time for medical evaluation was 3’26” and 3’19”, respectively. If the two reported severe TBIs, which an intensive care unit was on field, were excluded from the average, the mean time for medical evaluation goes down to 2’43” for players with concussion and 2’54” for replaced players after TBI. In comparison with 2014 FIFA World Cup, there was an increasing of time for player assessment on 2017 Brazilian Football Championship.

Many sports, including basketball, rugby, American football and ice hockey, have formal and feasible protocols for concussion. According to their rules, the athletes can be replaced to be evaluated on sideline and, according to protocol tests, they can return to play afterwards. Despite standardized protocols for concussion in football endorsed by FIFA, the main concern that arises from recent studies, including this one, is that there is a gap between these formal protocols and the sideline medical assessment in practice. In addition, the recognition of injury and assessment always occur in a time-pressured environment, requiring rapid disposition and decision making.[5] There is no doubt that the duration of this evaluation is very far from the ideal in order to preserve the player’s integrity.

Football rules forbid temporary replacement. This fact is probably the biggest challenge in the sport to deal with concussion assessment when compared with others team sports. It means that, in football environment, the medical professionals are pressured to perform concussion protocols on sideline as fast as possible because of a huge team disadvantage in the dynamic of the game without one player on the field. Our results confirm that the length of medical evaluation on sideline is not enough for an adequate performance. Consequently, the concussion and other potential neurologic injuries may be misdiagnosed, with a life threatening risk to the professional football player.

Finally, considering the previous studies already mentioned, this report suggests an alternative to the challenging issue of football’s replacement rules, that is: “the player with a concussion should be temporarily replaced to maintain the team with all players on the field and return to play after medical evaluation, if it is possible”. This will preserve the integrity of football players and provide an adequate sideline evaluation, respecting the dynamic and the environment of the sport.

## Data Availability

This is an observational study and all collected data from video analysis were systematically filled and are available with the research coordinator.

## Patient consent for publication

not required

